# Effects of esketamine on postoperative pain, anxiety, depression, sleep, and inflammation in patients undergoing cesarean section: A randomized controlled tria

**DOI:** 10.1101/2025.07.05.25330935

**Authors:** Yijun Wang, Xiaolu Lin, Xiang Zou, Renqin Zhang, Juanxia Xing, Li Zhang, Jie Shen, Xiaoliang Zhang, Daju Zhou, Junhua Zhang

**Author notes:** Corresponding author (JZ) or (DZ).

## Abstract

Postoperative pain is the most notable issue after cesarean section (CS). Contributing factors include hyperalgesia, anxiety, depression, sleep disorders, and inflammation. This study explored the effects of esketamine on pain, hyperalgesia, depression, anxiety, sleep disorders, and inflammation after CS. This randomized, double-blind, controlled trial enrolled single-term pregnant women scheduled for elective CS. Participants were randomly included in the esketamine group (group E: intravenous esketamine 0.5 mg/kg + sufentanil 4 μg/kg followed by patient-controlled intravenous analgesia [PCIA] with esketamine 0.5 mg/kg) or the control group (C: normal saline + sufentanil 4 μg/kg PCIA). The primary outcome was the maximum pain numerical rating scale (NRS) score within 24 h postoperatively. Secondary outcomes included pain NRS scores for moving incision, visceral, and rest incision pain at 0–6 h, 6–12 h and 12–24 h; pressure pain threshold and tolerance at 30 min and 24 h postoperatively; PCIA drug consumption and number of compressions; time to first PCIA compression; serum C-reactive protein (CRP) at 24 h; incidence of drug-related side effects; and rates of anxiety, depression, and sleep disorders on postoperative day 2. Ninety-eight women were randomly included in group E (n=50) or C (n=48). Group E had significantly lower maximum NRS pain scores within 24 h (5 [4–5] vs. 6 [5–6], P=0.000) and relieved rest incision, visceral, and moving incision pain at all time points. The PCIA compression was significantly delayed, and CRP levels, as well as the incidence of postoperative depression, anxiety, and sleep disorders, were lower in group E. There were no statistically significant differences in hyperalgesia or side effects. Intravenous esketamine effectively reduces postoperative pain, psychological disorders, and inflammation after CS.

This study was registered in the Chinese Clinical Trial Registry with registration number ChiCTR2300078310.

## Introduction

Pain is the most notable issue following cesarean section (CS) [1]. The incidence of acute moderate to severe postoperative pain after CS is as high as 68% [2], and chronic pain occurs in approximately 25.5% of cases [3]. Pain significantly affects postoperative recovery, mental state, breastfeeding, and the mother-infant relationship [4]. Patient-controlled intravenous analgesia (PCIA) is a commonly used analgesic method in clinical practice. However, high-dose opioids are associated with side effects such as constipation, nausea and vomiting, and respiratory depression, and they may also lead to tolerance, addiction, and hyperalgesia [5]. Therefore, there is an urgent need to explore new multimodal postoperative analgesic methods to relieve postoperative pain following CS.

Inflammatory reactions caused by tissue injury not only lead to inflammatory pain but also activate nerve terminals to generate action potentials that are transmitted to N-methyl-D-aspartic acid (NMDA) receptors in the spinal cord through the dorsal root ganglia. These signals activate specific brain areas to decrease the pain threshold, resulting in hyperalgesia [6, 7]. Moreover, pain has a bidirectional relationship with postoperative depression, anxiety, and sleep disorders [8]. Therefore, inflammation, hyperalgesia, anxiety, depression, and sleep disorders are important factors that aggravate pain after CS.

Esketamine is a novel analgesic with a non-competitive antagonistic effect on NMDA receptors, which may alleviate hyperalgesia. It also exhibits anti-inflammatory properties and has been shown to improve postpartum depression and sleep disorders [9–13]. However, studies on its use in CS are limited. Therefore, this study explored the effect of esketamine on postoperative pain after CS, including its impact on inflammation, hyperalgesia, anxiety, depression, and sleep disorders.

## Methods

This randomized, double blind, placebo-controlled trial was approved by the Chongqing University Fuling Hospital ethics committee (2023CDFSFLYYEC-064) and registered in the Chinese Clinical Trial Registry (registration number: ChiCTR2300078310). This study was conducted in Chongqing university Fuling Hospital. All procedures were performed in accordance with the Declaration of Helsinki guidelines (2024 edition) and followed the Consolidated Standards of Reporting Trials (CONSORT) reporting guidelines (2025 edition) (Fig. 1). All participants were informed of the trial details and provided written informed consent.

**Fig 1.**
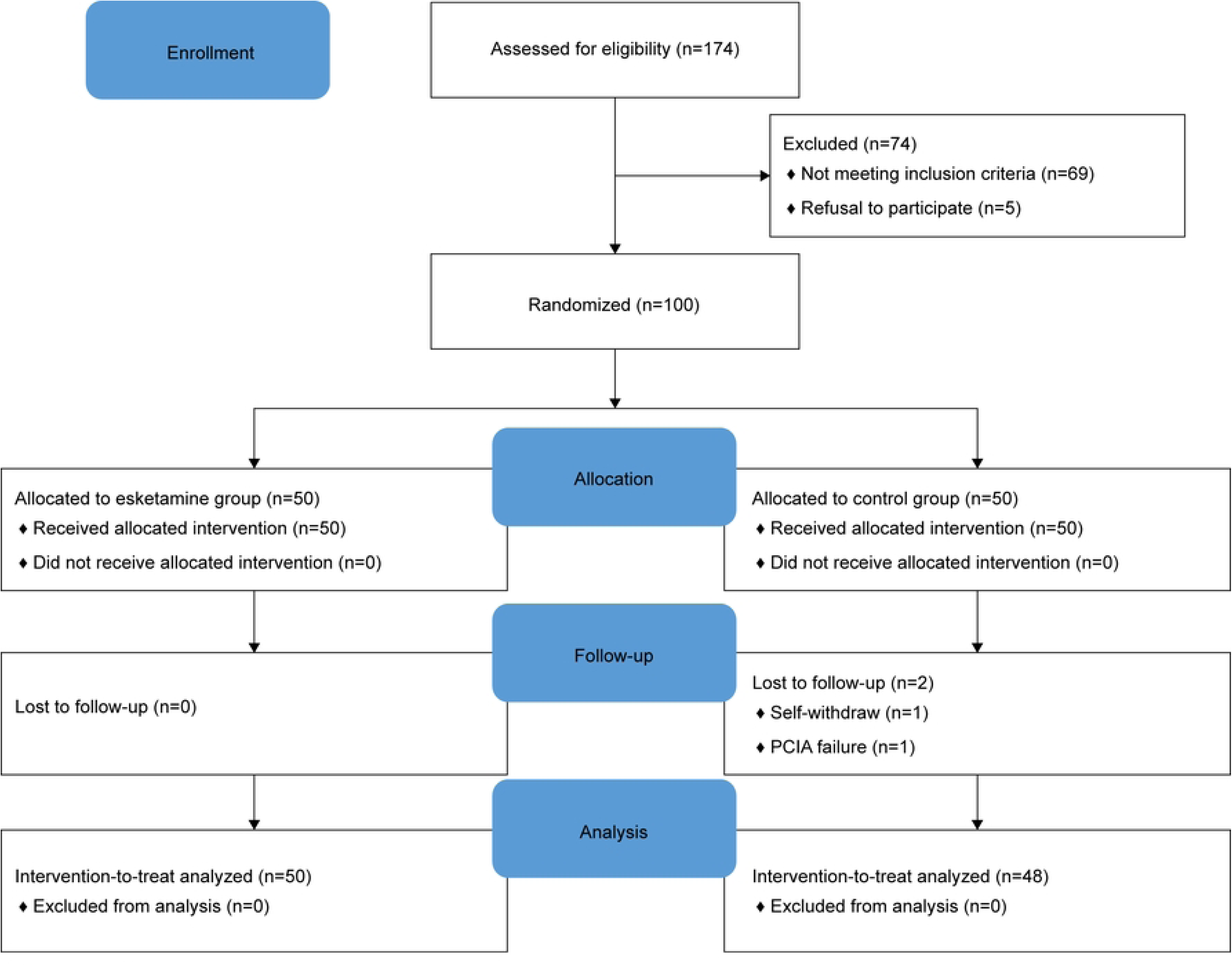
CONSORT Flowchart of the Study.

### Inclusion criteria

Included were women aged 20–45 years, with singleton full-term pregnancies, who underwent elective CS under combined spinal-epidural anesthesia, had American Society of Anesthesiologists (ASA) Physical Status Class II–III, and voluntarily participated after providing signed informed consent.

### Exclusion criteria

We excluded those with contraindications to CS, such as intrauterine stillbirth, fetal malformation, maternal intolerance to surgery, and fetal distress; contraindications to combined spinal-epidural anesthesia, such as coagulation dysfunction and central nervous system disorders; contraindications to any study drugs, including esketamine and sufentanil; history of alcoholism or chronic opioid, hormone, or anti-inflammatory/analgesic drug use; allergy to the study drugs; or inability or unwillingness to cooperate or provide informed consent.

### Randomization and masking

A simple randomization method was used. Random numbers were generated in a 1:1 ratio by an independent researcher using SPSS version 26.0 (IBM Corp, Armonk, NY). Another researcher, who was not involved in the study or data analysis, assigned the sequence and placed the random numbers in light-tight, sealed numbered envelopes. Participants were randomly included in the esketamine group (group E) or the control group (group C). A researcher not involved in the data collection and analysis prepared the study drugs and PCIA according to the random number allocation. All drugs and PCIA were labeled only as "investigational drugs" to mask their identity. The envelopes were re-secured after completion of the intervention. The anesthesiologists, participants, data analysts, and outcome assessors were all blinded to group assignments.

### Preoperative data collection

Informed consent was obtained 1 day before the CS. Demographic data and comorbidities were recorded. The Pittsburgh Sleep Quality Index was used to assess the sleep status in the previous month. The Athens Insomnia Scale was used to evaluate postoperative sleep quality on day 2. The Edinburgh Postnatal Depression Scale and Generalized Anxiety Disorder scale were used to respectively evaluate depression and anxiety preoperatively and on postoperative day 2.

A digital algometer pain diagnostic gauge (WAGNER, FDIX25 WAGNER INTERNATIONAL AG, USA [unit: kgf]) was used to test the pressure pain threshold (PPT) and pressure pain tolerance (PTO). The algometer was applied vertically to the skin at each test point, and pressure was applied slowly and uniformly. PPT was defined as the pressure at which pain was first perceived, and PTO was the pressure at which pain became intolerable (Fig. 1). Three test points, spaced 2 cm apart, were marked on the dominant forearm. The average values of the three measurements were recorded (Fig. 2).

**Fig 2.**
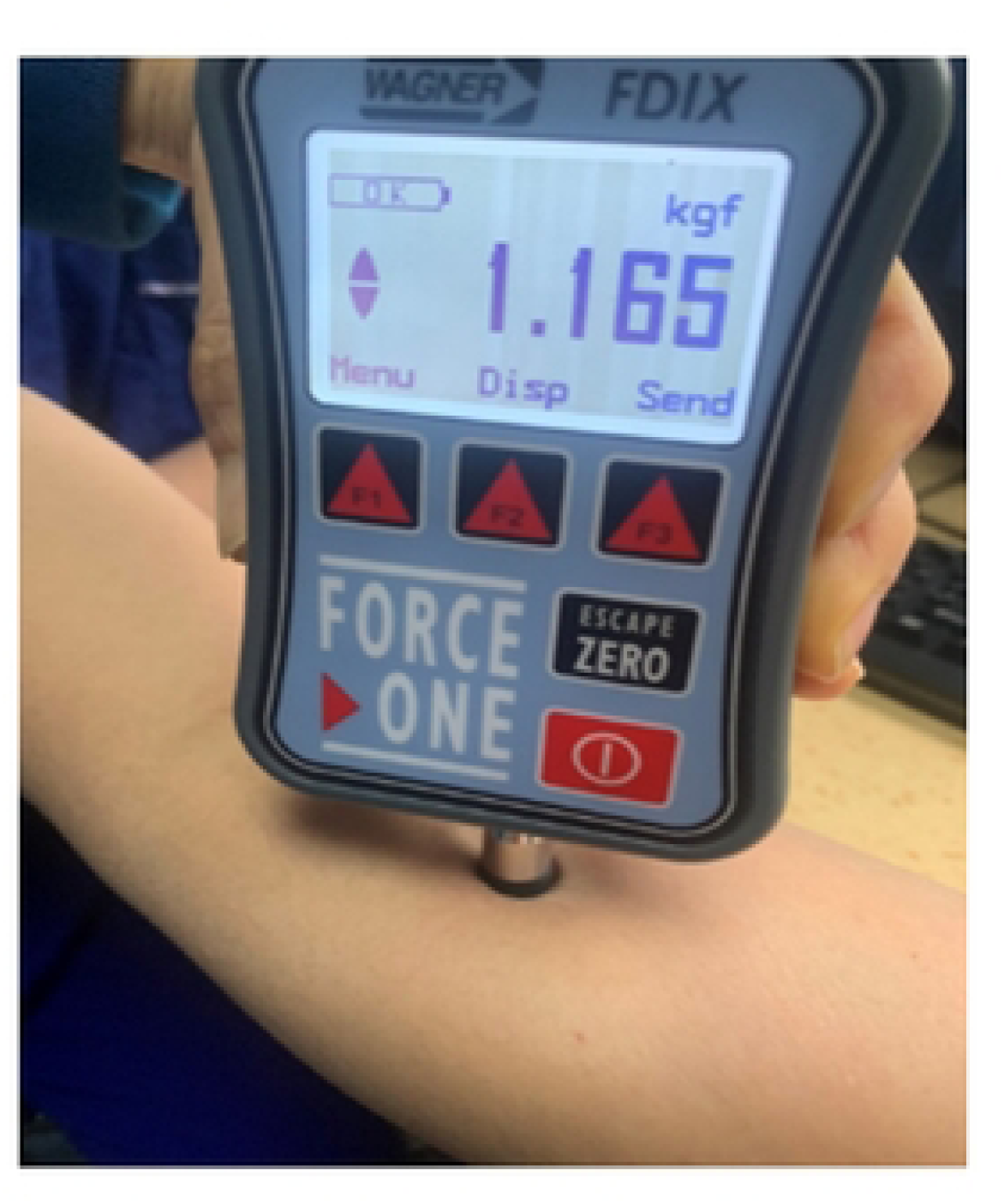
Digital Algometer Pain Diagnostic Gauge.

**Fig 3.**
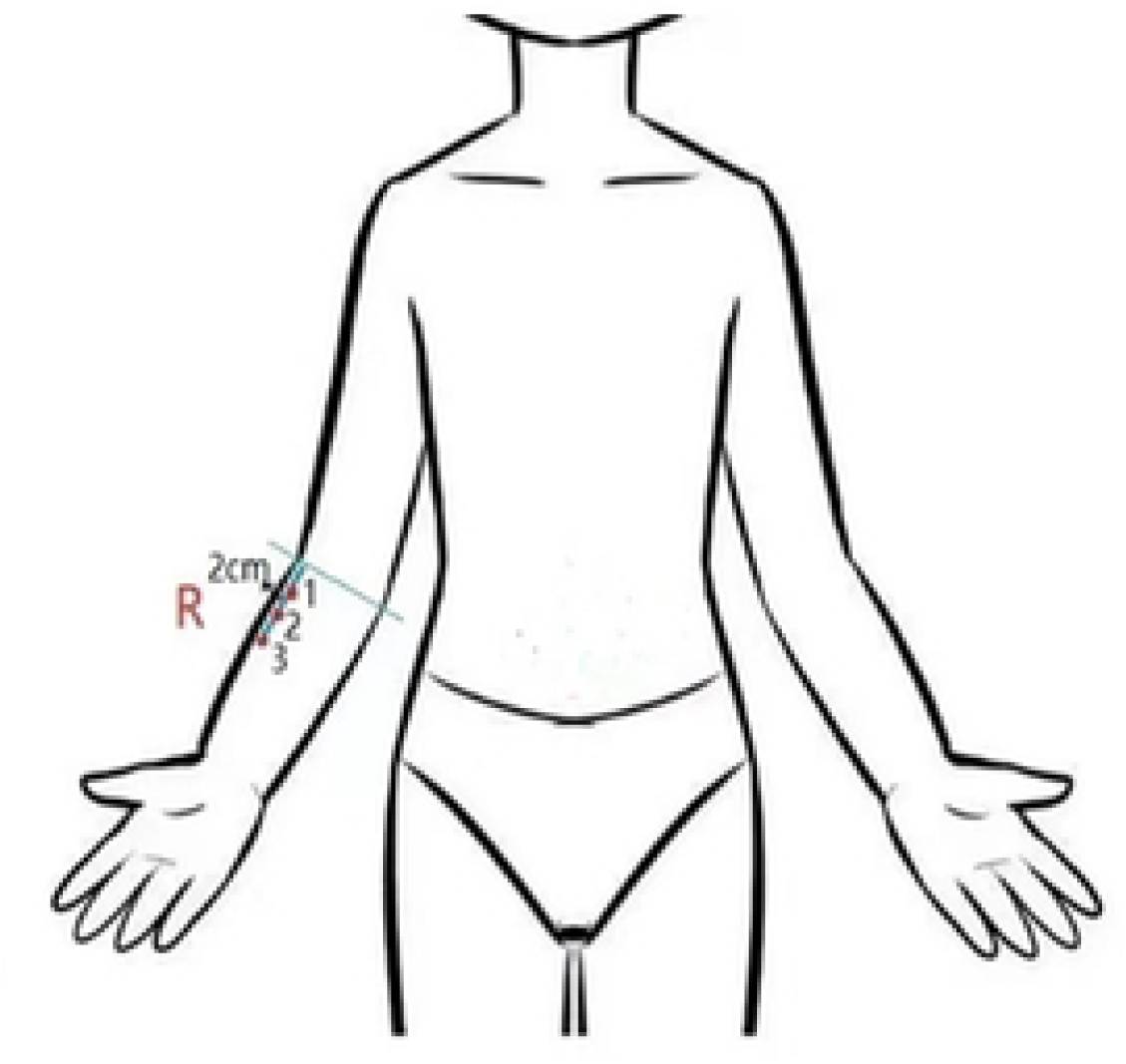
Test Points for the Pressure Pain Threshold and Pressure Pain Tolerance Measurements.

### Anesthesia and surgery method

After admission to the operating room, the participants were continuously monitored for vital signs including electrocardiography, pulse oxygen saturation, non-invasive blood pressure, and respiratory rate. A peripheral venous channel input was opened using Ringer’s solution at 500 ml/h. Combined spinal-epidural anesthesia was performed using a median puncture approach at the L3–4 spinal segment. In the subarachnoid space, 1.5 ml of 1% ropivacaine + 1.5 ml of 10% glucose injection, totaling 3 ml, was administered. A 4 cm epidural catheter was then inserted cephalad. The anesthesia level was adjusted to T6. If spinal anesthesia failed to reach T6, 5 ml of 2% lidocaine was injected epidurally. Norepinephrine was administered via infusion to maintain the mean arterial pressure at ≥80% or ≥60 mmHg. If the heart rate dropped below 50 bpm, 0.3 mg atropine was intravenously administered. If oxygen saturation was <90% or there was respiratory depression, 60% oxygen was administered via a mask. The CS was performed using a Pfannenstiel incision with peritoneal closure. After clamping the umbilical cord, oxytocin 10 IU was intravenously infused. At the end of the operation, 0.25 mg intravenous palonosetron (Hengrui Pharmaceutical Co., LTD., Jiangsu, China) was provided to prevent postoperative nausea and vomiting.

### Intervention

In group E, immediately after umbilical cord clamping, 0.5 mg/kg esketamine (Hengrui Pharmaceutical Co., LTD., Jiangsu, China) was slowly injected intravenously for approximately 1 min. A PCIA pump containing 4μg/kg sufentanil (Yichang Renfu Pharmaceutical Co., LTD., Hubei, China), 0.5 mg/kg esketamine, and 0.25 mg palonosetron, diluted with normal saline to 200 ml, was connected at the end of the surgery.

In Group C, immediately after umbilical cord clamping, intravenous injection of the same amount of normal saline was administered. The PCIA pump contained 4 μg/kg sufentanil and 0.25 mg palonosetron, also diluted with normal saline to 200 ml.

The PCIA parameter settings included: initial dose, 0 ml; constant infusion dose, 2 ml/h; bolus dose, 3 ml/time; locking time, 15 min; and maximum dose per h, 14 ml. If the PCIA dose reached the maximum, and the patient reported significant pain (numeric rating scale [NRS] score ≥4), 50 mg tramadol was injected intravenously.

### Outcomes

The primary outcome was the maximum NRS pain score within 24 h postoperatively. The secondary outcomes were the pain NRS scores for moving incision, visceral, and rest incision pain at 0–6 h, 6–12 h, and 12–24 h; time of the first PCIA compression, number of compressions (NOC), and total PCIA drug consumption during each time interval; PPT and PTO at 30 minutes and 24 hours postoperatively; serum C-reactive protein (CRP) concentration and incidence of drug-related side effects at 24 hours postoperatively; and incidence of anxiety, depression, and sleep disorders on postoperative day 2.

### Sample size calculation

This was a prospective trial. Based on data from a pilot study, the expected maximum pain score at 24 h after CS was 6.5±0.9 in the control group and 4.0±1.0 after prophylactic esketamine administration. We calculated that 45 participants would be needed in each group according to the 1:1 parallel control difference test, using SPSS sample size calculation software, test level of 5%, and test efficiency of 90%. Considering 10% loss to follow-up, the final sample size was set at 50 participants per group, for a total of 100 participants.

### Statistical analysis

Continuous variables that conformed to the normal distribution are expressed as mean ± standard deviation (̂x ± s), while those with a non-normal distribution are expressed as median (interquartile range [IQR]) [M (Q25, Q75)]. Categorical variables are expressed as absolute values and frequencies. Data conforming to a normal distribution were compared using independent sample t-tests. Non-normally distributed variables (e.g., NRS pain scores) were analyzed using non-parametric tests (Mann– Whitney U tests). Fisher’s exact probability method or chi-squared tests were used to compare differences in nausea and vomiting, dizziness, drowsiness, nightmares, and diplopia between the groups. SPSS version 26.0 was used for the statistical analyses. P≤0.05 was set as the threshold of statistical significance.

## Results

We screened 174 pregnant women, of whom, 100 were included and randomly assigned to the esketamine group (n=50) or control group (n=50). The first and final participant was included on December 12, 2023 and April 18, 2025 respectively. There was no unblinding throughout the study period. In group C, two participants dropped out: 1 withdrew owing to PCIA failure 3 h postoperatively and 1 voluntarily withdrew 8 h after surgery. The remaining 98 participants completed the study procedure and data analysis.

At baseline, the age of the participants was significantly lower in group E than in group C, and group C included more cases of gestational hypothyroidism. There were no significant differences in terms of gestational age, demographic characteristics (weight, height, and body mass index), history of CS, or other complications (Table 1).

**Table 1.**
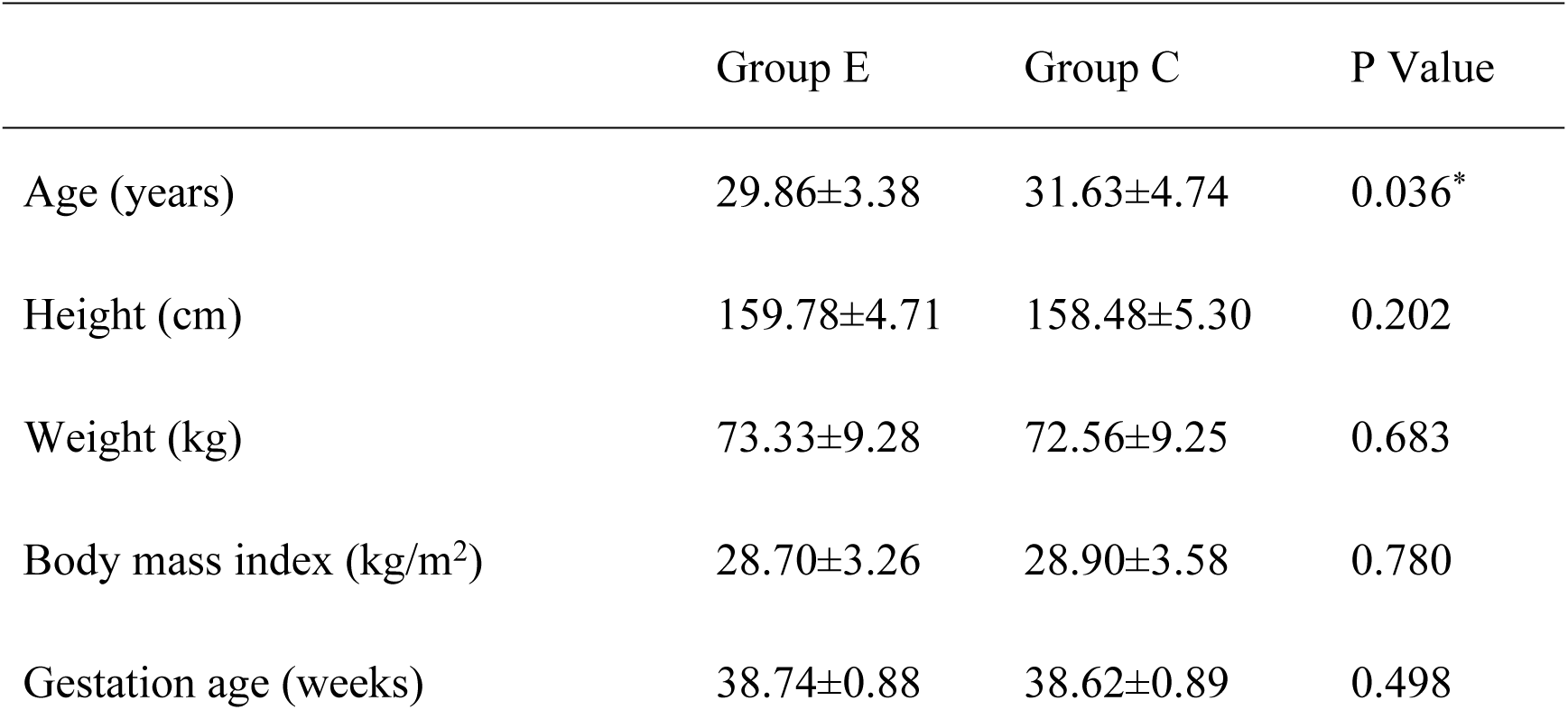
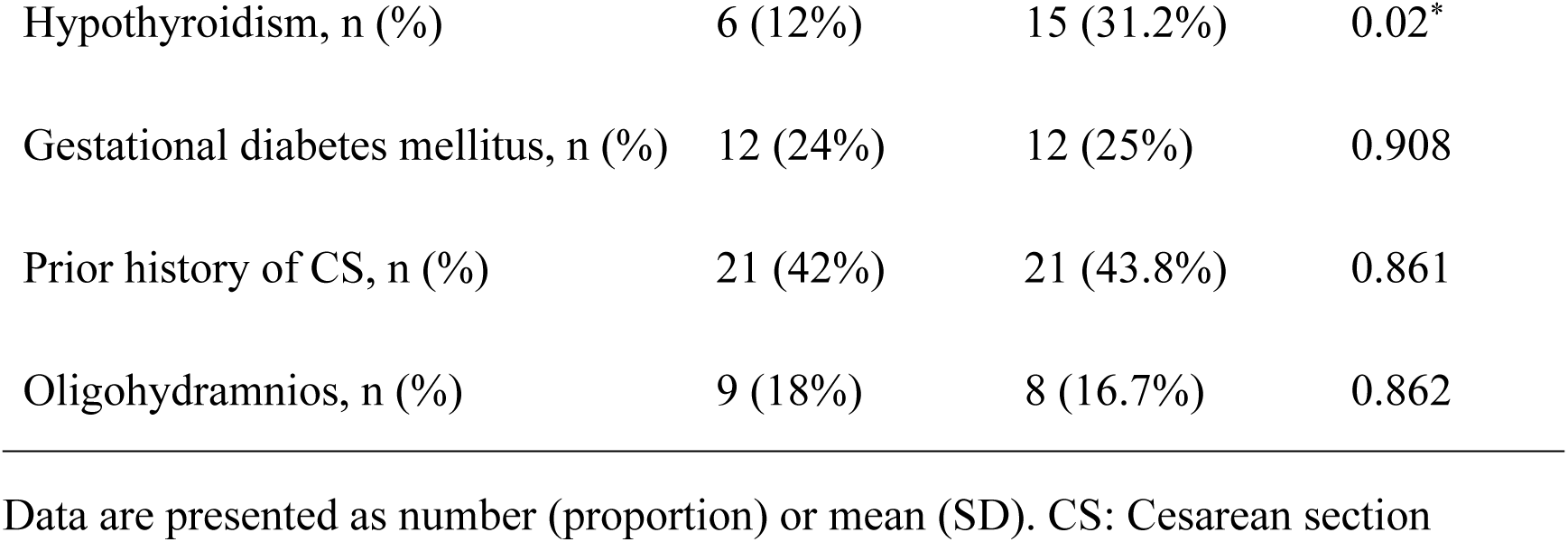
Demographic Characteristics of the Participants in the Two Groups.

The maximum NRS pain score within 24 h after surgery was lower in group E than in group C (median [IQR]: 5 [4–5] vs. 6 [5–6], P=0.000). In addition, the maximum NRS scores for moving incision, visceral, and rest incision pain were significantly lower in group E than in group C at any period.

Rest incision pain:

- 0-6 h: 1 [0–1.25] vs. 1 [1–2], P=0.000
- 6–12 h: 1 [1–2] vs. 2 [1.25–2], P=0.000
- 12–24 h: 2 [1–2] vs. 2 [1, 3], P=0.000

Moving incision pain:

- 0–6 h: 3 [2–3] vs. 4 [3–4], P = 0.000
- 6–12 h: 4 [3–5] vs. 5 [4–5], P = 0.000
- 12–24 h: 4 [4–5] vs. 5 [4–5], P = 0.016

Visceral pain:

- 0–6 h: 2 [2–3] vs. 3 [3–4], P=0.000
- 6–12 h: 3 (2–4) vs. 4 [3–5], P=0.003
- 12–24 h: 4 [2.75–4.25] vs. 5 [4–5. 75], P=0.000 (Fig. 4).

**Fig 4.**
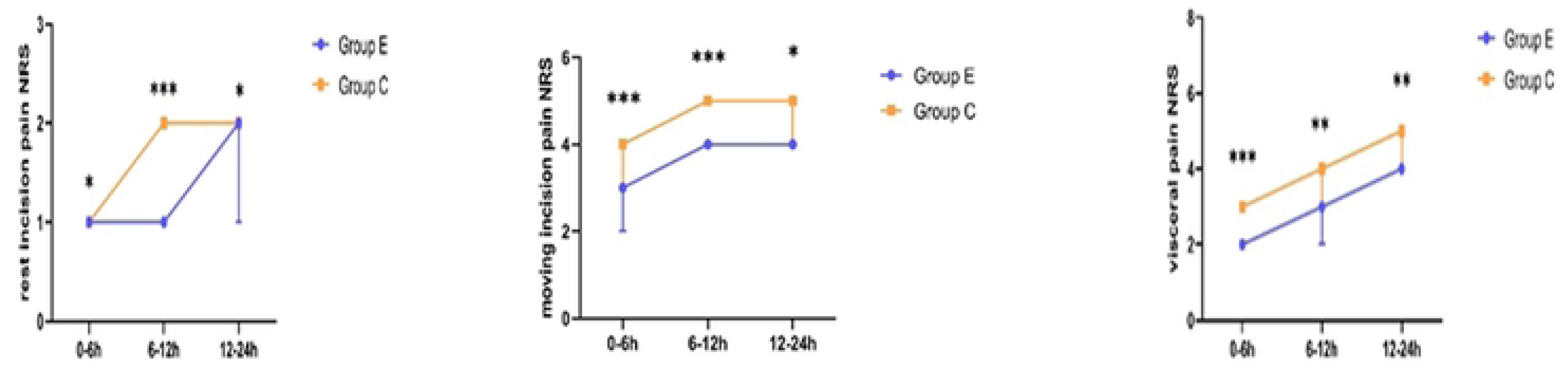
Comparison of NRS Scores for Rest Incision, Moving Incision, and Visceral Pain Between Group E and Group C at Each Postoperative Time Point. NRS: Numeric Rating Scale

The time to first PCIA compression was significantly delayed in group E over that in group C. However, there were no differences between the groups in NOC and total PCIA consumption at any time point (Table 2).

**Table 2.**
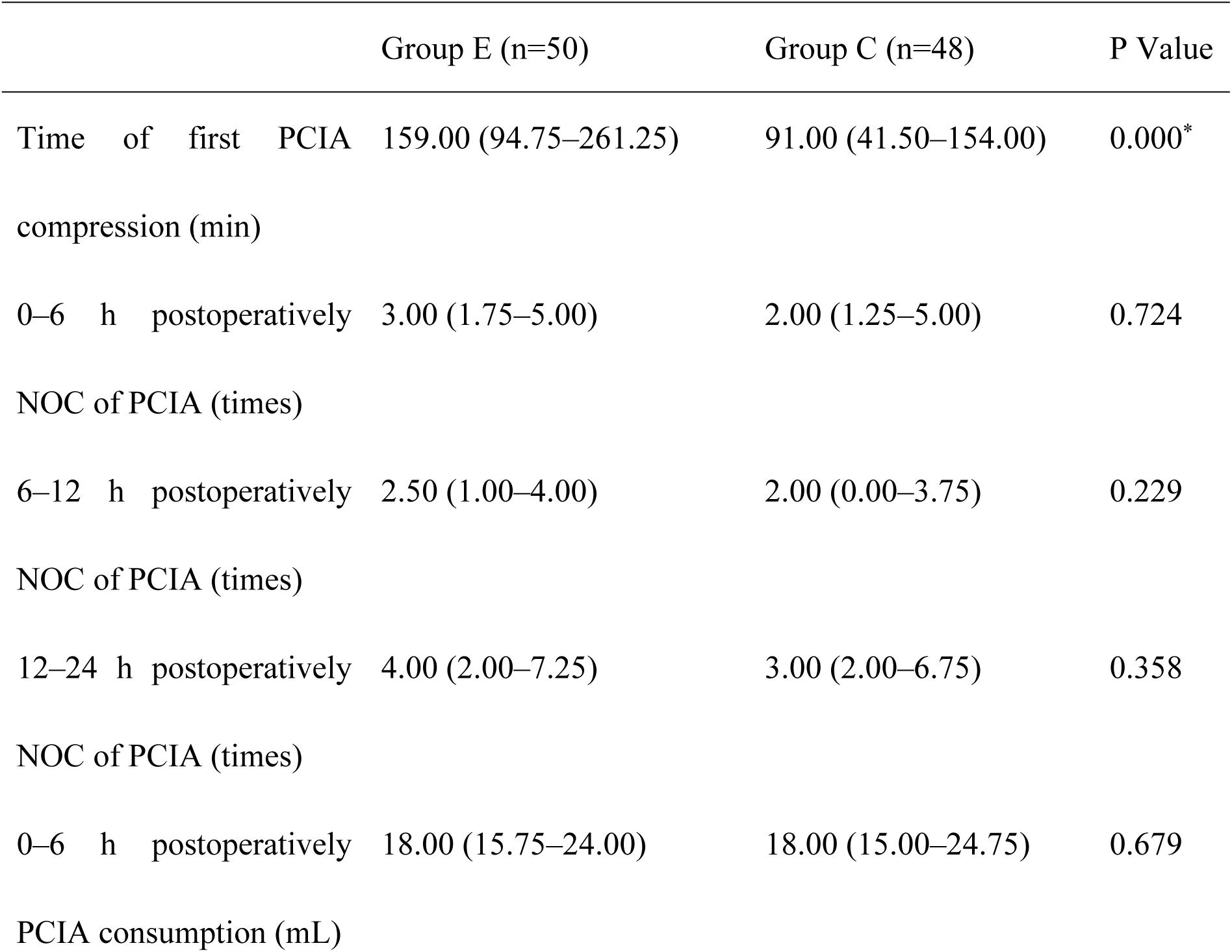
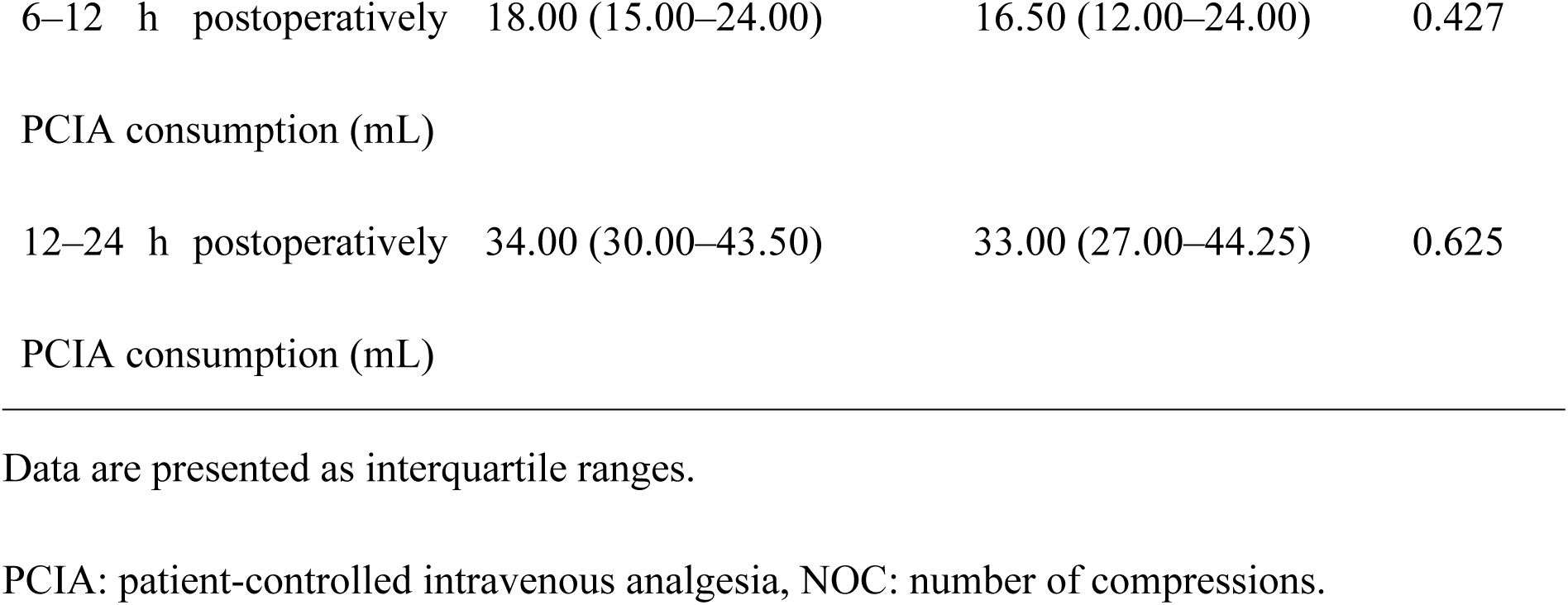
Usage of the PCIA Pump.

The preoperative and postoperative PPT and PTO values showed no significant difference between the groups.

PPT (kgf):

- T0: 2.1 [1.62–2.4] vs. 2.2 [1.59–2.86], P=0.677
- T1: 2.18 [1.76–2.6] vs. 2.33 [1.79–2.95], P=0.440
- T2 2.03 [1.6–2.83] vs. 2.15 [1.8–2.8], P = 0.376)

PTO (kgf):

- T0: 4 [3.2–5.05] vs. 4.25 [3.36–5.2], P = 0.513
- T1: 4.29 [3.1–5.3] vs. 4.4 [3.38–5.56], P = 0.558
- T2: 3.88 [2.62–4.83] vs. 3.85 [3.33–4.92], P = 0.272 (Fig. 5)

**Fig 5.**
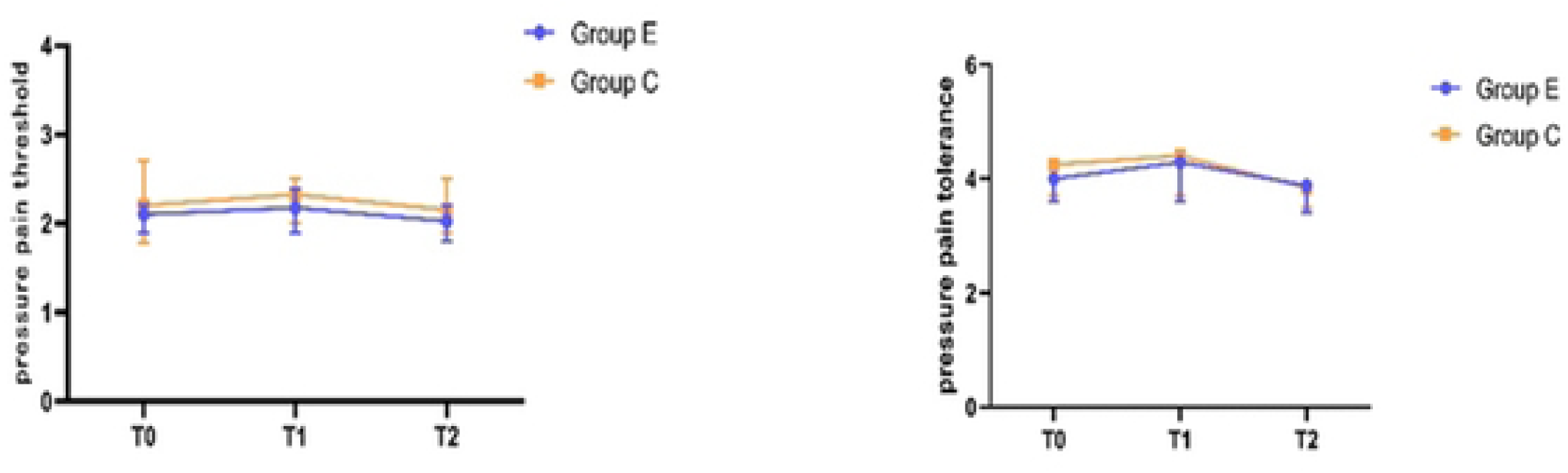
Comparison of Preoperative and Postoperative PPT and PTO Between the Groups. PPT: pressure pain threshold, PTO: pressure pain tolerance

The preoperative sleep, anxiety, and depression evaluations were not significantly different between the groups. However, on postoperative day 2, the incidence of sleep disorders, postpartum depression, and anxiety was significantly lower in group E (Table 3).

**Table 3.** Comparison of Anxiety, Depression, and Sleep Disorders Between the Groups.

|  | Group E (n=50) | Group C (n=48) | P Value |
| --- | --- | --- | --- |
| Preoperative sleep quality |  |  |  |
| Good | 42 (84%) | 39 (81.3%) | 0.567 |
| Relatively good | 8 (16%) | 4 (8.3%) |  |
| Average | 0 (0%) | 5 (10.4%) |  |
| Poor | 0 (0%) | 0 (0%) |  |
| Postoperative sleep quality |  |  |  |
| Suspected insomnia | 0 (0%) | 11 (22.9%) | 0.000 * |
| Insomnia | 1 (2%) | 8 (16.7%) |  |
| No insomnia | 49 (98%) | 29 (60.4%) |  |
| Preoperative anxiety |  |  |  |
| No | 43 (86%) | 37 (77.1%) | 0.228 |
| Mild | 7 (14%) | 9 (18.8%) |  |
| Moderate | 0 (0%) | 1 (2.1%) |  |
| Severe | 0 (0%) | 1 (2.1%) |  |
| Postoperative anxiety |  |  |  |
| No | 49 (98%) | 39 (81.3%) | 0.006 * |
| Mild | 1 (1%) | 9 (18.7%) |  |
| Moderate | 0 (0%) | 0 (0%) |  |
| Severe | 0 (0%) | 0 (0%) |  |
| Preoperative depression |  |  |  |
| No | 37 (74%) | 37 (77.1%) | 0.868 |
| Mild | 13 (26%) | 8 (16.7%) |  |
| Depression | 0 (0%) | 3(6.2%) |  |
| Postoperative depression |  |  |  |
| No | 48 (96%) | 40 (83.3%) | 0.038 * |
| Mild | 2 (4%) | 7 (14.6%) |  |
| Depression | 0 (0%) | 1 (2.1%) |  |

Data are presented as numbers (proportion)

**Table 4.** Comparison of the Incidence of Drug-related Side Effects Between the Groups.

|  | Group E (n=50) | Group C (n=48) | P Value |
| --- | --- | --- | --- |
| Nausea and vomiting | 8 (16%) | 2 (4.2%) | 0.092 |
| Dizziness | 4 (8%) | 3 (6.3%) | 1.000 |
| Drowsiness | 5 (10%) | 2 (4.2%) | 0.436 |
| Nightmare | 5 (10%) | 2 (4.2%) | 0.436 |
| Diplopia | 1 (2%) | 0 (0%) | 1.000 |

Data are presented as numbers (proportion)

There was no difference in preoperative serum CRP concentrations (3.70±1.40 vs. 4.08±1.78 mg/L, P=0.237). However, the postoperative serum CRP concentration was significantly higher in group C than in group E (73.92±28.20 vs. 59.32±17.24 mg/L, P=0.003) (Fig. 6).

**Fig 6.**
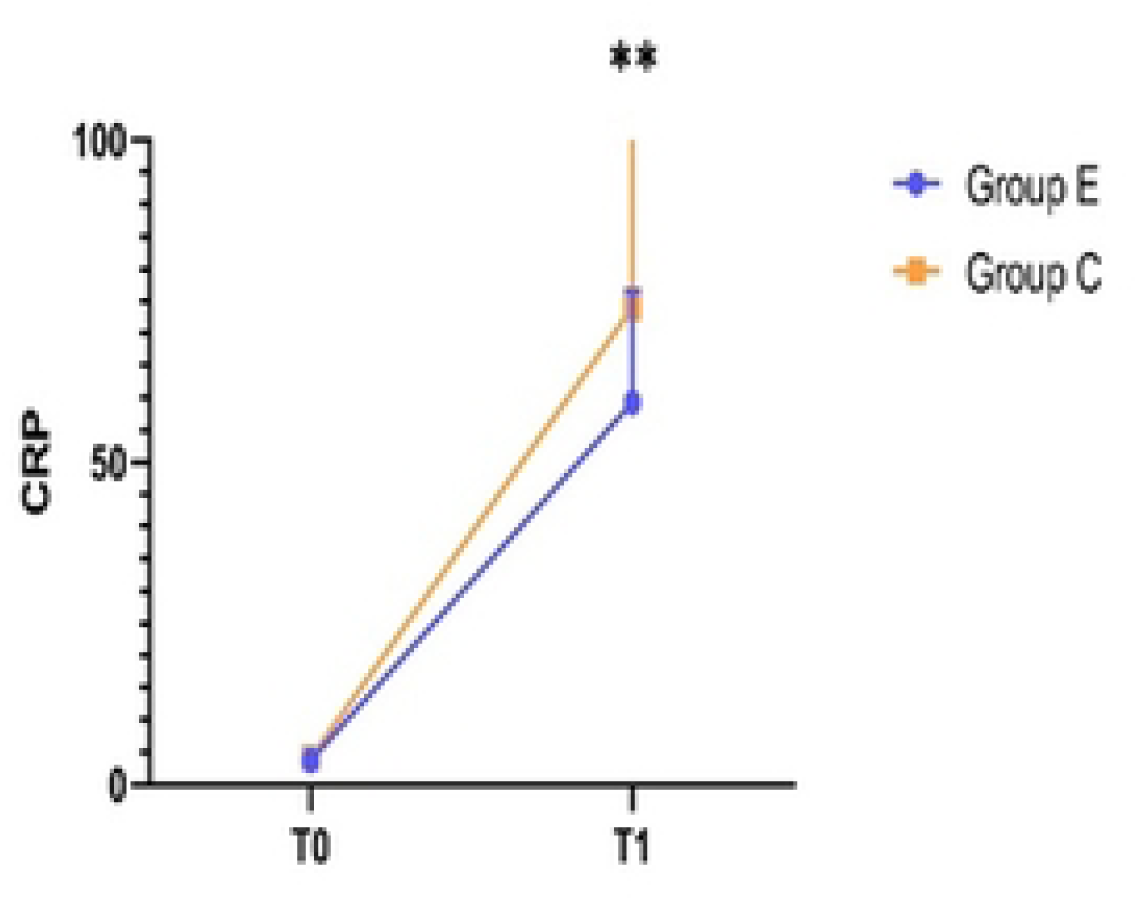
Comparison of Serum CRP Concentrations Between the Groups. CRP: C-reactive protein

The incidence of side effects (dizziness, nausea and vomiting, drowsiness, nightmares, and diplopia) was not significantly different between the groups.

## Discussion

In this clinical trial, we found that the combined application of esketamine effectively alleviated postoperative pain within 24 h after CS and had an anti-inflammatory effect; it also reduced the incidence of anxiety and depression and improved sleep quality on postoperative day 2. However, esketamine had no significant effect on hyperalgesia.

There are close bidirectional interactions between sleep disorders, anxiety, depression, and pain. Sleep disorders are known to exacerbate pain by disrupting the inflammatory system, aggravating inflammatory responses, and interfering with pain regulation and processing. However, they also heighten pain perception and reduce pain tolerance [14]. Additionally, pain and sleep disorders can significantly increase the risk of anxiety and depression and even lead to suicide [15].

As an optical isomer of ketamine, esketamine blocks pain signal conduction through non-competitive antagonism of NMDA receptors to produce analgesic effects. It also regulates the glutamatergic system, which crucially influences pain, anxiety, depression, and sleep, thereby improving postoperative anxiety, depression, and sleep quality [16]. Our findings on esketamine’s postoperative analgesia and antidepressant effects are consistent with those of previous research[17–20]. As far as we know, this is the first study to report the effects of esketamine on anxiety and sleep disorders after CS.

Tissue injury during surgery induces an inflammatory response that contributes to postoperative pain [21]. This process involves the activation of NMDA receptors in the dorsal horn of the spinal cord through the dorsal root ganglia, leading to neuroinflammation and activating sensory neurons. The result is a lowered pain threshold and increased pain sensitivity, causing hyperalgesia [22]. Therefore, we hypothesized that esketamine may be effective in alleviating post-CS hyperalgesia owing to its non-competitive NMDA receptor antagonistic effect. Although Ren et al. found that esketamine effectively relieved hyperalgesia after thyroidectomy [23], we found no significant effect on hyperalgesia in our study; we and other research teams have previously reported this finding [24, 25]. Hyperalgesia after thyroidectomy may result not only from surgical tissue injury but also from opioid-induced hyperalgesia. Therefore, we hypothesized that esketamine would be more effective in relieving opioid-induced hyperalgesia than that caused by tissue injury.

At baseline, the incidence of pregnant women with preoperative hypothyroidism was significantly higher in group C than in group E. Although reports on the interaction between hypothyroidism and pain are few, Ørstavik et al. suggested that hypothyroidism, which is one of the causes of painful neuropathy, can affect large- and small-fiber function, thereby changing the body’s pain sensitivity [26]. Therefore, in this study, the higher NRS pain scores observed in group C may reflect the confounding effect of more patients with hypothyroidism.

In addition, the mean age of participants in group E was lower than that in group C. Wang and Worly discovered in a retrospective study that older pregnant women (aged 36–45 years) had a higher demand for analgesic drugs than younger pregnant women, whereas younger pregnant women exhibited a greater incidence of severe pain [27]. Similarly, Lokeshwar et al. suggested that younger maternal age is associated with more severe postoperative pain, possibly due to anxiety [28]. In our study, despite the younger average age in group E, their NRS pain scores were lower than those in group C, indicating a powerful analgesic effect of esketamine.

There are some limitations to this study. First, this was a single-center study, which may limit the generalizability of the findings across different regions and populations. Second, given concerns regarding participant recruitment owing to the declining birth rate in China, we set the dropout rate at 10% during the sample size calculation. Although the final dropout rate was less than 10%, the overall sample size was relatively small. Future multicenter studies with larger sample sizes are required. Finally, this study was limited to the short-term effects of esketamine after CS. The long-term effects of esketamine on postoperative chronic pain, anxiety, depression, and sleep conditions remain unknown.

## Conclusion

Esketamine effectively relieved postoperative pain after CS, reduced anxiety and depression, improved sleep quality, and exerted anti-inflammatory effects. However, it did not affect postoperative hyperalgesia.

## Data Availability

Individual participant data are available from the sponsor upon reasonable request through email

## Acknowledgments

None.

## Data availability statement

Individual participant data are available from the sponsor upon reasonable request through email.

